# Systemic ELOVL6 activity predicts survival and represents a modifiable target of ALS

**DOI:** 10.64898/2026.04.30.26352060

**Authors:** Andrés Jiménez-Zúñiga, Gorka Fernández-Eulate, José I. Ruíz-Sanz, Jose Luis Zúñiga-Elizari, Maddi Garciandia, Javier Riancho, Raul Domínguez, Ahmad Al Khleifat, Monica Zufiría, Sonia Alonso-Martin, Roberto Fernández-Torrón, Juan J. Poza-Aldea, Jon Ondaro, Juan B. Espinal, Gonzalo González-Chinchón, Amaia Martinez-Arroyo, Miren Zulaica, Maria B. Ruiz-Larrea, Ammar Al-Chalabi, Maialen Sagartzazu, Ian J. Holt, Mónica Povedano, Adolfo López de Munain, Gorka Gereñu, Francisco J. Gil-Bea

## Abstract

**Background:** Amyotrophic lateral sclerosis (ALS) is characterized by profound metabolic reprogramming, yet the lack of biomarkers for specific druggable targets remains a major hurdle for precision medicine. We hypothesized that peripheral lipid biosynthetic signatures could serve as both prognostic indicators and a roadmap for identifying novel disease-modifying targets.

**Methods:** We assessed serum fatty acid (FA) metabolic pathways in two independent longitudinal cohorts (n = 37 and n = 38) using high-dimensional CoxBoost modeling. Primary outcomes were survival and functional staging milestones, including non-invasive ventilation and gastrostomy. The biological relevance of the identified candidate was further assessed through correlation with plasma neurofilament light-chain (NfL) levels. Causality and therapeutic potential were validated in Drosophila melanogaster models of TDP-43 proteinopathy via genetic ablation and pharmacological inhibition.

**Results:** Our multi-parametric model, comprising two clinical variables and the estimated ELOVL6 (elongation of very long-chain fatty acids protein 6) activity, demonstrated robust prognostic accuracy (Uno’s C 0.69) across both cohorts; ELOVL6 activity served as a strong independent predictor of mortality and functional decline. Notably, high ELOVL6 activity significantly correlated with elevated plasma NfL levels (p < 0.01), linking peripheral elongation imbalances to central axonal damage. In Drosophila, ELOVL6 overactivation was identified as a conserved pathological consequence of TDP-43 dysfunction, characterized by an increased C18:0/C16:0 ratio in both loss-of-function and gain-of-function models. Inhibition of ELOVL6, either genetically or pharmacologically, rescued neuromuscular junction integrity, prolonged survival, and significantly reduced pathological TDP-43 phosphorylation in glial models.

**Conclusion:** These findings position ELOVL6 as a promising modifiable metabolic node with potential for disease-modifying intervention in ALS. Beyond its potential utility for identifying high-risk metabolic profiles and assisting in prognostic counseling, ELOVL6 bridges systemic lipid dysregulation with TDP-43 proteinopathy. Targeting this pathway offers a dual opportunity: as a biological marker to supplement clinical staging and as a druggable enzymatic target to ameliorate motor neuron degeneration.

**HIGHLIGHTS:**

- Systemic ELOVL6 activity is a robust independent predictor of ALS survival.
- High ELOVL6 levels correlate with plasma NfL and functional decline.
- Inhibition of ELOVL6 rescues NMJ integrity and survival in *Drosophila* models.
- Pharmacological targeting of ELOVL6 reduces glial TDP-43 phosphorylation.
- ELOVL6 represents a druggable metabolic node linking lipids to proteinopathy.

## Background

Amyotrophic lateral sclerosis (ALS) is a devastating and heterogeneous neurodegenerative disorder characterized by the progressive loss of motor neurons in the motor cortex, brainstem, and spinal cord (Brown and Al-Chalabi, 2017). Despite decades of research, diagnosis still relies on clinical and electrophysiological criteria, leading to delays of about one year from onset (Ittner et al., 2015). More critically, the translational success rate for disease-modifying therapies has been remarkably poor (Riancho et al., 2019). This failure is increasingly attributed to the profound biological heterogeneity of the patient population and an incomplete understanding of the early molecular drivers that precede irreversible neuronal degeneration (Brown and Al-Chalabi, 2017; Riancho et al., 2019). Consequently, there is an overarching need to identify biomarkers that do not merely reflect the consequence of neurodegeneration, but rather represent actionable pathogenic nodes suitable for therapeutic intervention.

In recent years, the paradigm of ALS has shifted from a purely proteocentric view to one that encompasses a systemic metabolic collapse (Bouteloup et al., 2009; Jésus et al., 2018). A hallmark of this collapse is a state of hypermetabolism, characterized by increased resting energy expenditure and a paradoxical shift in substrate utilization (Jésus et al., 2018). While healthy neural tissue relies heavily on glucose, the ALS-affected system undergoes metabolic reprogramming toward increased fatty acid (FA) oxidation and a depletion of adipose tissue reserves (Fergani et al., 2007). Accumulating evidence suggests that dysregulation of the lipidome is not a secondary bystander effect but a central player in disease pathogenesis (Palamiuc et al., 2015; Dobrowolny et al., 2018). In clinical cohorts, higher levels of circulating triglycerides and cholesterol have been associated with prolonged survival, suggesting that lipid homeostasis provides a vital neuroprotective buffer against the energetic deficit characteristic of ALS (Dupuis et al., 2008; Dorst et al., 2011; Paganoni et al., 2011). However, the relevance of lipids extends beyond their role as energy substrates; their specific saturation profile dictates cellular toxicity. An imbalance towards saturated and long-chain fatty acids, also referred to as lipotoxicity, has been recognized as a stressor to which neurons are uniquely vulnerable (Guttenplan et al., 2021; Yang et al., 2025). While plasma neurofilament light chain (NfL) has emerged as the gold-standard biomarker for monitoring axonal injury (Khalil et al., 2024), its utility remains primarily non-disease-specific. Although NfL has proven valuable for both diagnosis and prognosis, it reflects the consequence of neurodegeneration rather than a modifiable cause, limiting its ability to guide metabolic interventions. Thus, shifting the focus to lipid metabolism offers a unique opportunity to identify “first-in-class” targets. Unlike NfL, enzymatic nodes represent upstream drivers that can be pharmacologically modulated to prevent neurodegeneration.

Recent large-scale transcriptomic and lipidomic analyses have further reinforced this metabolic paradigm, demonstrating that a loss of neuronal polyunsaturated fatty acids (PUFAs) and a concomitant shift toward lipid saturation are conserved features of ALS/FTD across multiple species (Giblin et al., 2025). Crucially, while increasing PUFA levels is neuroprotective, the accumulation of long-chain saturated species—primarily driven by imbalances in the desaturation and elongation machinery—appears to sensitize motor neurons to degeneration (Giblin et al., 2025). Indeed, the clinical relevance of FA elongation is underscored by pathogenic variants in elongases such as ELOVL1, ELOVL4, and ELOVL5, which are associated with diverse neurodegenerative disorders (Ferrero et al., 2025). Notably, ELOVL1 has been identified as a potential therapeutic target in X-linked adrenoleukodystrophy and reactive astrocyte-mediated toxicity, where reducing long-chain saturated FAs mitigates neuronal death (Sassa et al., 2014; Boyd et al., 2021; Come et al., 2021).

We previously demonstrated a systemic trend toward longer-chain fatty acids in ALS patients, suggesting an early dysregulation of the elongation pathway (Fernandez-Eulate et al., 2020). In the present study, we hypothesized that the peripheral lipidomic signature of ALS patients could be interrogated to reveal specific dysregulated enzymatic nodes that drive disease progression. By profiling serum FAs across two independent longitudinal cohorts and utilizing high-dimensional CoxBoost modeling, we identified that the activity of ELOVL6 (elongation of very long-chain fatty acids protein 6), the rate-limiting enzyme in the elongation of C16 to C18 FAs, serves as a strong, independent predictor of survival and functional staging milestones.

To transition from prognostic correlation to mechanistic causality, we utilized *Drosophila melanogaster* models to investigate whether ELOVL6 overactivation constitutes a modifiable driver of ALS pathology. We demonstrate that the inhibition of ELOVL6, via both genetic ablation and selective pharmacological intervention, remarkably ameliorates neuromuscular junction (NMJ) denervation, locomotor deficits, and the pathological phosphorylation of TDP-43. By integrating clinical lipidomic interrogation with functional *in vivo* validation, we position ELOVL6 as a novel disease-modifying target, providing a metabolic roadmap for therapeutic intervention in ALS.

## Methods

### Study design and participant recruitment

#### Training cohort

Patients with suspected motor neuron disease evaluated at the Motor Neuron Disease Unit of Bellvitge University Hospital (Barcelona, Spain) were recruited between May 2014 and May 2017 (n = 37). Inclusion criteria were: i) fulfillment of El Escorial criteria for probable or definite ALS (spinal or bulbar onset), and ii) blood sampling performed within one year of diagnosis. Clinical follow-up data were collected until December 12, 2019, with a total follow-up time of 78.2 months.

#### Validation cohort

An independent validation cohort (n = 38) was generated by consecutively recruiting patients fulfilling El Escorial criteria for probable or definite ALS from three referral centers in Northern Spain (Donostia University Hospital, Araba University Hospital, and Marqués de Valdecilla University Hospital) between April 2011 and May 2017. Data collection concluded on January 10, 2020, with a total follow-up time of 106.8 months.

In both cohorts, electromyography (EMG) was performed to fulfill El Escorial criteria. Baseline clinical variables including age, sex, diagnostic delay, and site of onset were collected for all participants. The percentage of theoretical Forced Vital Capacity (%FVC) and Body Mass Index (BMI) were available for the training cohort.

While the validation cohort exhibited a significantly shorter diagnostic delay compared to the training cohort (p = 0.01) (Table 1), suggesting a potentially more progressive phenotype, this diversity provided a rigorous test for the generalizability of the identified metabolic biomarkers.

**Table 1.** Demographic and clinical characteristics of patients with ALS at study in each cohort.

|  | Training cohort | Validating cohort | p-value |
| --- | --- | --- | --- |
| Number of patients | 37 | 38 | -- |
| Age at sampling (years)<sup>a</sup> | 61.6 ± 13.0 | 63.4 ± 12.1 | 0.55<sup>d</sup> |
| Sex (male/female) | 20/17 | 26/12 | 0.20<sup>e</sup> |
| Cause (sporadic/familial) | 37/0 | 34/4 | 0.04<sup>e</sup> |
| Type of onset (spinal/bulbar) | 28/9 | 33/5 | 0.21<sup>e</sup> |
| %FVC<sup>a</sup> | 64.3 ± 24.5 | -- | -- |
| ALSFRS-R slope<sup>a,b,c</sup> | 0.74 ± 0.56 | -- | -- |
| FTD (yes/no) | 4/33 | 2/36 | 0.38<sup>e</sup> |
| Diagnostic delay (days)<sup>a</sup> | 404.5 ± 233.3 | 274.0 ± 181.7 | 0.01<sup>d</sup> |
| BMI (kg)<sup>a</sup> | 25.8 ± 3.7 | -- | -- |
| Treatment (statin/antidiabetic) | 7/1 | 5/1 | 0.82<sup>e</sup> |
| Follow-up (months) | 78.2 | 106.8 | -- |

<sup>a</sup>Values given as mean ± standard deviation.
<sup>b</sup>ALSFRS-R slope is given as the decline of score per month and is calculated using the following formula: (48 minus ALSFRS-R score) / (date of ALSFRS-R score minus date of onset).
<sup>c</sup>ALSFRS-R slope is available from 20 patients.
<sup>d</sup>Student's t test.
<sup>e</sup>Chi-square test.

#### Clinical outcomes and censoring

To assess survival and disease progression, a minimum observation window of 2.5 years from sampling was established, based on the reported median survival of 2–3 years for ALS. The primary endpoint was all-cause mortality. The secondary endpoint was defined as the placement of non-invasive ventilation (NIV) or percutaneous endoscopic gastrostomy (PEG), serving as proxies for King’s clinical stage 4.

For survival analysis, patients alive at the end of the follow-up were censored. For the secondary endpoint analysis, patients with pre-existing NIV or PEG at the time of sampling were excluded. To avoid confounding by indication (e.g., refusal of life-sustaining measures), patients who died without receiving NIV or PEG despite clinical indication were excluded from the secondary analysis. Patients alive and free of NIV/PEG at the end of follow-up were censored.

### Serum FA profiling

Total lipids were extracted from serum using a chloroform/methanol (2:1, v/v) mixture. FAs were transmethylated following the protocol described before (Lepage and Roy, 1986). Fatty acid methyl esters (FAMEs) were analyzed by gas chromatography coupled with a flame ionization detector (GC-FID) using an HP5890 Series II system (Hewlett Packard), as previously described (Ruiz-Sanz et al., 2018). GC-FID was selected for its high reproducibility and ability to precise double-bond localization, which is essential for assessing elongation and desaturation pathways. FA concentrations were expressed as a percentage of total FAs. To estimate the enzymatic activity of desaturases and elongases, product-to-precursor ratios were calculated as detailed in Table S1.

Specifically, systemic ELOVL6 activity was defined as the composite sum of the following product-to-precursor ratios: C14:0/C12:0, C16:0/C14:0, C18:0/C16:0, and C18:1n-7/C16:1n-7. While the C18:0/C16:0 ratio serves as the primary enzymatic indicator in tissue-specific models, this composite sum was utilized in serum to provide a more stable and robust systemic signature for clinical prognostic modelling.

### Quantification of plasma Neurofilament Light Chain (NfL)

Plasma NfL concentrations were quantified using the single molecule array (Simoa) NF-light™ assay on an HD-1 Analyzer (Quanterix, Lexington, MA) at the Maurice Wohl Clinical Neuroscience Institute (London, UK). Samples were randomized, blinded, and analyzed in duplicate using a single batch of reagents to minimize variation. The intra-assay and inter-assay coefficients of variation were 4.8% and 10.5%, respectively.

### Drosophila models and experimental procedures

#### Fly stocks and genetics

*Drosophila melanogaster* stocks were maintained under standard conditions at 22°C for stocking, with 70% humidity and a 12h:12h light/dark cycle. To model ALS-associated proteinopathy, the GAL4/UAS system was utilized for tissue-specific expression. For muscle-specific knockdown of *Tbph* (the *Drosophila* ortholog of *TARDBP*), the *Mef2-GAL4* driver (VDRC stock #15549) was employed to generate two distinct models: a moderate model (iTbphM, using VDRC stock #38377/GD6943) characterized by reduced lifespan, and a severe model (iTbphS, using VDRC stock #104401/pkk108354) exhibiting 100% pharate adult lethality. Experiments with these models were done at 22°C. Genetic inhibition of ELOVL6 was achieved using a *UAS-Baldspot-RNAi* line (targeting the *Drosophila* ELOVL6 ortholog) obtained from the BDSC repository (Stock #35321). Control flies were generated by crossing the respective drivers with a line carrying a *pVal20* vector to knockdown *Gal4* (UAS-iGal4; BDSC #35784). To assess glial toxicity, a gain-of-function model (Glia-hTDP43) was established by crossing the glial-specific *REPO-GAL4* driver (BDSC #7415) with a *UAS-hTDP-43* line (BDSC #79587). Experiments with glial model were done at 26°C.

#### Pharmacological treatment

For pharmacological validation, a selective indoledione ELOVL6 inhibitor was synthesized as previously described(Shimamura et al., 2009). The compound was dissolved in vehicle (DMSO) and incorporated into the standard cornmeal medium at a final concentration determined of 10 µM to provide optimal efficacy. Treatment was initiated from the larval stages or upon eclosion, depending on the assay requirements. Control groups received an equivalent volume of vehicle.

#### Lifespan analysis

For longevity tests, 50 adult female flies per genotype were housed in groups of five. Survival was monitored every 2 days, and food was replaced at least once per week.

#### Pharate survival

For the severe iTbphS model, “pharate adult survival” was quantified as the percentage of adult flies that successfully eclosed from the pupal case relative to the total number of pupae in each vial.

#### Locomotor activity

Climbing assays were performed on specific days (e.g., days 7 and 14). Groups of five flies were placed in a tube with a mark at 8 cm; the number of flies crossing this line within 10 seconds was recorded over three trials.

#### Immunofluorescence and synaptic analysis

Adult flies were anesthetized with CO_2_-enriched air. Brains and thoraces were isolated, immersed in 70% EtOH for 1 minute, and placed in PBS for dissection. Samples were fixed with 4% PFA for 30 minutes at room temperature. To expose the flight muscles, thoraces were immersed in liquid nitrogen for 10 seconds, transferred to cold PBS, and sectioned longitudinally using a feather double-edge razor blade (Aname). Following fixation, brains and thoraces were washed three times in PBS containing 0.5% Triton X-100 (PBS-T) and incubated in blocking solution (5% BSA and 0.02% NaN_3_ in PBS-T) for 30 minutes at room temperature. Samples were then incubated overnight at 4°C with the following primary antibodies: rabbit anti-hTDP-43 (1:100; Cell Signaling), rat anti-p-hTDP-43 (Ser409/410) (1:200; BioLegend), mouse anti-REPO (1:10; DSHB), mouse anti-Bruchpilot (nc82) (1:100; DSHB), and rabbit anti-HRP (1:100; Jackson Immuno-Research). After four 20-minute washes with PBS-T, samples were incubated overnight at 4°C with secondary antibodies conjugated to Alexa Fluor Plus fluorophores (Invitrogen, 1:1000). To visualize nuclei and muscle structure, DAPI (ThermoFisher, 1:500) and Alexa Fluor™ 647 phalloidin (Invitrogen, 1:1000) were added to the brains and thoraces, respectively. Samples were mounted using ProLong™ Diamond Antifade Mountant (Invitrogen). Confocal images were acquired with a Zeiss LS900 microscope using Z-stack scanning of whole-mount brains or flight muscles.

#### Drosophila lipidomics

Lipid extraction from whole flies or dissected tissues (thoracic muscle/brain) was performed using the Folch method. FA methyl esters were generated and analyzed by GC-FID as described above for human serum samples.

### Statistical modeling and variable selection via CoxBoost

To address the high dimensionality of the dataset relative to the sample size, we employed a component-wise boosting approach for Cox proportional hazards models using the **CoxBoost** R package. Unlike gradient boosting methods that rely on loss function gradients, CoxBoost adapts an offset-based boosting strategy for parameter estimation, as described by Tutz and Binder (Tutz and Binder, 2007). This approach results in sparse model fits analogous to Lasso-like regularization, effectively performing simultaneous variable selection and parameter estimation by shrinking non-informative coefficients to zero.

#### Optimization Strategy

The model architecture was optimized via two key tuning parameters: the penalty for partial likelihood estimation and the number of boosting steps.

1. Penalty Parameter: The optimal penalty value was determined using the optimCoxBoostPenalty function to maximize the coarsened partial likelihood.
2. Boosting Steps: Using the optimized penalty, the number of boosting steps was selected via leave-one-out cross-validation (LOOCV) using the cv.CoxBoost function. Following established recommendations for likelihood-based boosting performance, we ensured the search space allowed for an optimal number of steps greater than 50 to prevent premature stopping.

Once the hyperparameters were fixed, the final model was fitted using the CoxBoost function to identify the subset of variables with non-zero coefficients.

#### Evaluation of model performance

We assessed the prognostic accuracy of the model in both the training and independent validation cohorts using three complementary metrics:

1. Inverse Probability of Censoring (IPC) Weighted Uno’s C-Statistic. To assess global discrimination, we calculated Uno’s C-statistic. While analogous to Harrell’s concordance index (C-index), Uno’s C employs inverse-probability-of-censoring (IPC) weights to correct for the potential bias introduced when censoring is dependent on the covariate distribution (Uno et al., 2011). This makes it a more robust estimator for survival data with varying follow-up times. To address the absence of %FVC in the external validation cohort, we performed validation using the ELOVL6 activity as a standalone predictor. This allowed us to assess whether the metabolic signature provides prognostic value independently of standard respiratory metrics. Resampling Strategy: To generate robust confidence intervals, we trained the model on 75% of randomly selected samples from the training cohort. We then performed 50 resampling iterations on either the remaining 25% hold-out test set (internal validation) or the complete external validation cohort. The median values and 95% confidence intervals (CI) of Uno’s C-statistic were computed using the survAUC R package.
2. Time-Dependent Incident/Dynamic AUC. To evaluate the discriminative ability of the model at specific time points, we calculated the time-dependent Area Under the ROC Curve (AUC). Given the heterogeneous progression rates characteristic of ALS, the incident/dynamic AUC provides a more granular assessment of predictive power over time compared to global summary statistics (Heagerty and Zheng, 2005). Calculation: Incident/dynamic AUCs were calculated using the survivalROC and risksetROC R packages, implementing the nearest neighbor estimator method described by Bansal and Heagerty. Prediction Scores: To rigorously address the risk of overfitting inherent in high-dimensional datasets with limited sample sizes, we utilized a cross-validation framework to report all internal validation metrics. While leave-one-out cross-validation was employed for hyperparameter optimization (boosting steps and penalty), the final prognostic performance in the training cohort, including Uno’s C-statistic and time-dependent AUC, was derived using 5-fold cross-validation. This ensures that the reported internal validation performance reflects the model’s ability to generalize to unseen subjects within the cohort, providing a more conservative and robust estimate of its predictive power compared to standard training-set performance.
3. Log-Rank Test and Risk Stratification. To visualize clinical utility, patients were stratified into “Low-Risk” and “High-Risk” subgroups based on the median prediction score generated by the CoxBoost model. Survival Analysis: Survival distributions for the stratified groups were estimated using the Kaplan-Meier method and compared using the log-rank test (survival R package). Scoring Source: Similar to the AUC analysis, prediction scores for stratification were obtained via 5-fold cross-validation in the training cohort and from the median-performance model in the validation cohort. Note: While stratification provides a clear visual representation of risk divergence, we acknowledge that dichotomizing continuous risk scores ignores within-group heterogeneity; therefore, this metric was used as a complement to the continuous C-statistic and AUC measures.

### General statistics

For continuous variables (e.g., NfL levels, FA ratios), normality was assessed using the Shapiro-Wilk test. Group comparisons were performed using the Mann-Whitney U test or Student’s t-test as appropriate. Survival curves were compared using the log-rank test. A p-value < 0.05 was considered statistically significant. All analyses were performed using R statistical software.

## Results

### Identification of ELOVL6 Overactivation via High-Dimensional Modeling

Demographic and clinical profiles of the study cohorts are detailed in Table 1. In the training cohort (n = 37), 5 patients were censored at the end of the follow-up (December 12, 2019), with a median follow-up of 48.7 months (range: 32.4–53.6). The remaining 33 patients reached the primary endpoint (death), with a median survival of 17.0 months from sampling. In the validation cohort (n = 38), 8 patients were censored (January 10, 2020; median follow-up: 45.3 months), and 30 patients died, exhibiting a median survival of 13.2 months from sampling.

We observed specific alterations in FA profiles between ALS patients and healthy controls, although some inconsistencies were noted between cohorts (Tables S1–S3). For instance, while a reduction in the relative abundance of C20:3n-6 was robust across cohorts (Table S1), its modest magnitude limits its utility as a diagnostic marker, particularly given the availability of superior alternatives such as NfL. Conversely, we noted prominent shifts in enzymatic indices, specifically in ELOVL6-mediated elongation (C12–C16) and SCD1-mediated desaturation (Table S2). Crucially, our primary objective was not merely to distinguish patients from controls, but to identify lipid signatures capable of predicting disease trajectory, as these are more likely to represent actionable disease modifiers.

To interrogate the metabolic landscape and identify robust predictors of disease trajectory, we applied CoxBoost proportional hazards modeling to the training cohort. We integrated 55 serum FAs variables with six established clinical predictors (age, sex, site of onset, %FVC, diagnostic delay, and BMI) (Table S4). After 100 boosting steps (Figure S1), the model converged on three non-zero coefficients: age, %FVC at sampling, and systemic ELOVL6 enzymatic activity. We defined overall ELOVL6 activity as the composite sum of specific product-to-precursor ratios: C14:0/C12:0, C16:0/C14:0, C18:0/C16:0, and C18:1n-7/C16:1n-7 (Table S1, Figure S2). Univariate Cox regression confirmed the significance of these variables (Figure 1A), identifying elevated ELOVL6 activity and older age as significant risk factors (HR > 1), whereas higher %FVC was protective (HR < 1).

**Figure 1.**
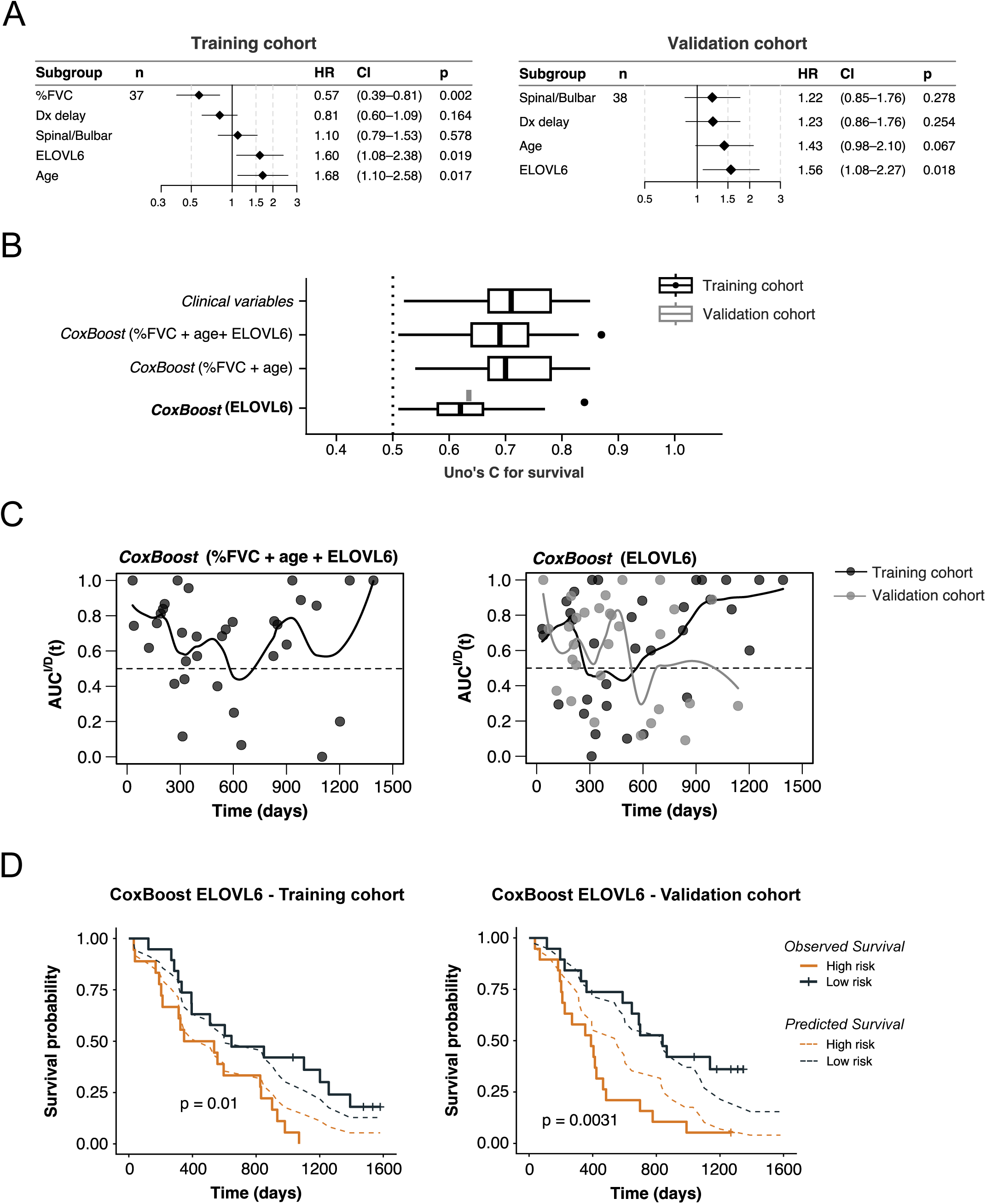
ELOVL6 activity is a robust independent predictor of ALS survival. **(A)** Univariate Cox proportional hazards regression analysis in the training cohort (n = 37). The forest plot displays Hazard Ratios (HR) for the variables selected by the high-dimensional CoxBoost model (Age at diagnosis, %FVC, and ELOVL6 activity) alongside standard clinical covariates (Diagnostic delay and Site of onset). **(B)** Predictive performance assessment using the Inverse Probability of Censoring (IPC)-weighted Uno’s C-statistic. Boxplots compare the prognostic accuracy of the Clinical model (Age + %FVC + Delay + Onset), the composite CoxBoost model (Age + %FVC + ELOVL6), the reduced model (Age + %FVC), and ELOVL6 activity alone in the training cohort (black). The predictive validity of ELOVL6 alone was confirmed in the independent validation cohort (n = 38, gray). **(C)** Time-dependent incident/dynamic Area Under the Curve [AUC(t)] analysis for survival. The plots illustrate the discriminative ability over time for the composite model (left) and ELOVL6 activity alone (right) across training and validation cohorts. **(D)** Kaplan-Meier survival analysis stratified by ELOVL6 risk profiles. Patients were classified as low-risk (< median, blue) or high-risk (≥ median, orange). Solid lines represent observed survival, while dashed lines indicate the model-predicted survival probability, demonstrating calibration. P-values derived from the log-rank test. Abbreviations: %FVC, percent forced vital capacity; ELOVL6, elongation of very long-chain fatty acids protein 6.

The multivariate model demonstrated strong prognostic accuracy, yielding an IPC-weighted Uno’s C statistic of 0.69 (95% CI: 0.50–0.82) in the training cohort. This performance was comparable to a model based exclusively on standard clinical variables (C = 0.71) and consistent with established risk scores such as the ENCALS prediction model (Figure 1B).

### ELOVL6 activity independently stratifies ALS mortality risk

To isolate the specific contribution of ELOVL6 as a potential pathogenic driver, we performed a sensitivity analysis by removing it from the multivariate model. The reduced model (age + %FVC) yielded a similar Uno’s C (0.70), while ELOVL6 alone maintained informative prognostic power (C = 0.62 in training; Figure 1B). To rigorously test the generalizability of this metabolic biomarker, we performed validation in an independent external cohort characterized by significant phenotypic heterogeneity, including a shorter diagnostic delay (p = 0.01) that likely reflects a more progressive disease state (Table 1). Crucially, as clinical respiratory metrics (%FVC) were unavailable for the validation cohort, we assessed ELOVL6 activity as a standalone predictor. Despite the absence of established clinical covariates and the more aggressive phenotype of this cohort, ELOVL6 activity alone demonstrated a robust and comparable prognostic accuracy (C = 0.64; Figure 1B). This confirms that the identified enzymatic signature provides independent prognostic value even when standard clinical markers are missing, highlighting its reliability as a versatile biological indicator of disease trajectory.

Time-dependent ROC analysis indicated that the standalone ELOVL6 model outperformed others in predicting 1-year survival (Figure 1C; Figure S3). Notably, higher ELOVL6 scores consistently stratified individuals at higher risk of early mortality across both cohorts. Consequently, the elevated ELOVL6 indices initially observed in the global case-control comparison (Table S2) are likely driven by the disproportionate weight of rapid progressors. This underscores the necessity of molecular stratification, as dysregulation of this elongation pathway is not a uniform feature across the entire patient population.

Kaplan-Meier analysis confirmed that patients classified as high-risk (ELOVL6 ≥ median) had significantly shorter survival compared to the low-risk group in both the training (p = 0.01) and validation (*p* = 0.0031) cohorts (Figure 1D). While the model showed good calibration in the training set, risk was slightly overestimated in the high-risk group of the validation set. However, the discriminative capacity remained robust: the ELOVL6-integrated model, the clinical-only model, and the reduced model all successfully differentiated high- vs. low-risk groups (*p* < 0.01; Figure S4). Furthermore, stratifying patients by ELOVL6 tertiles revealed a dose-dependent relationship, where patients in the highest tertile exhibited significantly reduced survival (training, *p* = 0.010; validation, *p* = 0.033).

### ELOVL6 overactivation mirrors functional decline and axonal injury

We next evaluated whether ELOVL6 activity correlated with functional disease milestones, specifically the time to NIV or PEG. In a combined dataset (n = 50), ELOVL6 activity demonstrated robust predictive ability for NIV (C = 0.68), PEG (C = 0.66), or the composite endpoint (C = 0.60) (Figure 2A). High ELOVL6 scores were significantly associated with an earlier requirement for NIV (*p* = 0.00025), PEG (*p* = 0.00053), or both (*p =* 0.0014) (Figure 2B).

**Figure 2.**
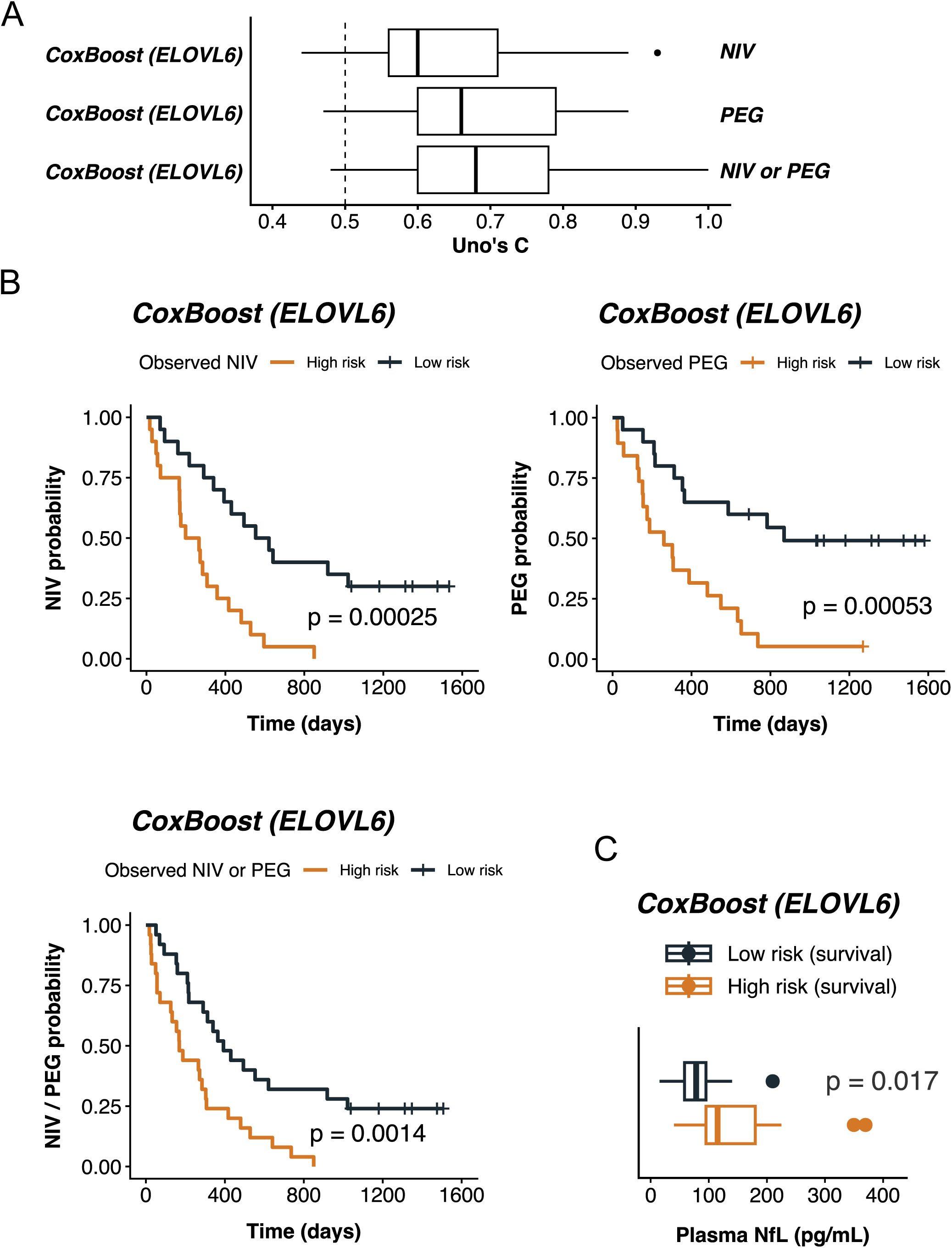
ELOVL6 activity predicts functional decline and mirrors axonal injury. **(A)** Prognostic accuracy for functional milestones. Boxplots display the Inverse Probability of Censoring (IPC)-weighted Uno’s C-statistics for predicting the requirement of non-invasive ventilation (NIV; n = 40), percutaneous endoscopic gastrostomy (PEG; n = 39), or the composite endpoint (n = 50) based on ELOVL6 activity in the combined study cohorts. **(B)** Kaplan-Meier analysis of disease progression. Patients were stratified into low-risk (dark blue) and high-risk (orange) groups based on median ELOVL6 activity scores. Log-rank tests demonstrate that high ELOVL6 activity is significantly associated with an earlier need for NIV, PEG, or the composite endpoint. **(C)** Association with markers of neurodegeneration. Boxplots compare plasma neurofilament light chain (NfL) levels in a patient subset (n = 35). Patients classified as high-risk by the ELOVL6 survival model exhibit significantly elevated baseline plasma NfL levels compared to the low-risk group (95-180 vs 60-100 pg/mL). Abbreviations: NIV, non-invasive ventilation; PEG, percutaneous endoscopic gastrostomy; NfL, neurofilament light chain.

Additionally, we examined plasma NfL levels. In a patient subset (n = 35), high NfL levels trended toward poorer survival (*p* = 0.017). Importantly, patients classified as high-risk by their ELOVL6 profile exhibited significantly elevated plasma NfL levels (Figure 2C). This correlation reinforces the biological relevance of ELOVL6, suggesting that its metabolic overactivation is closely tied to the rate of axonal degeneration and disease severity.

### Muscle-specific models: genetic and pharmacological inhibition rescues survival and NMJ integrity

To test the strategy of inhibiting ELOVL6 as a potential treatment for ALS, we performed functional experiments using *Drosophila melanogaster* models. We utilized two models, previously characterized by our group, in which the ortholog of *TARDBP* (*Tbph*) is knocked down specifically in muscle cells using the Mef2-GAL4 driver. In the moderate model (iTbphM), the knockdown results in a significantly reduced lifespan, while the more severe model (iTbphS) exhibits 100% pharate adult lethality, preventing the flies from emerging from their pupal stage (Figure 3A). Based on these backgrounds, we generated new models where the knockdown of *Baldspot* (the fly ELOVL6 ortholog) is combined with either the moderate (iTbphM-iBaldspot) or severe (iTbphS-iBaldspot) knockdown of *Tbph* in muscle cells (Figure 3B). These integrated models allowed us to evaluate the efficacy of the ELOVL6 inhibition strategy under conditions of varying disease severity.

**Figure 3.**
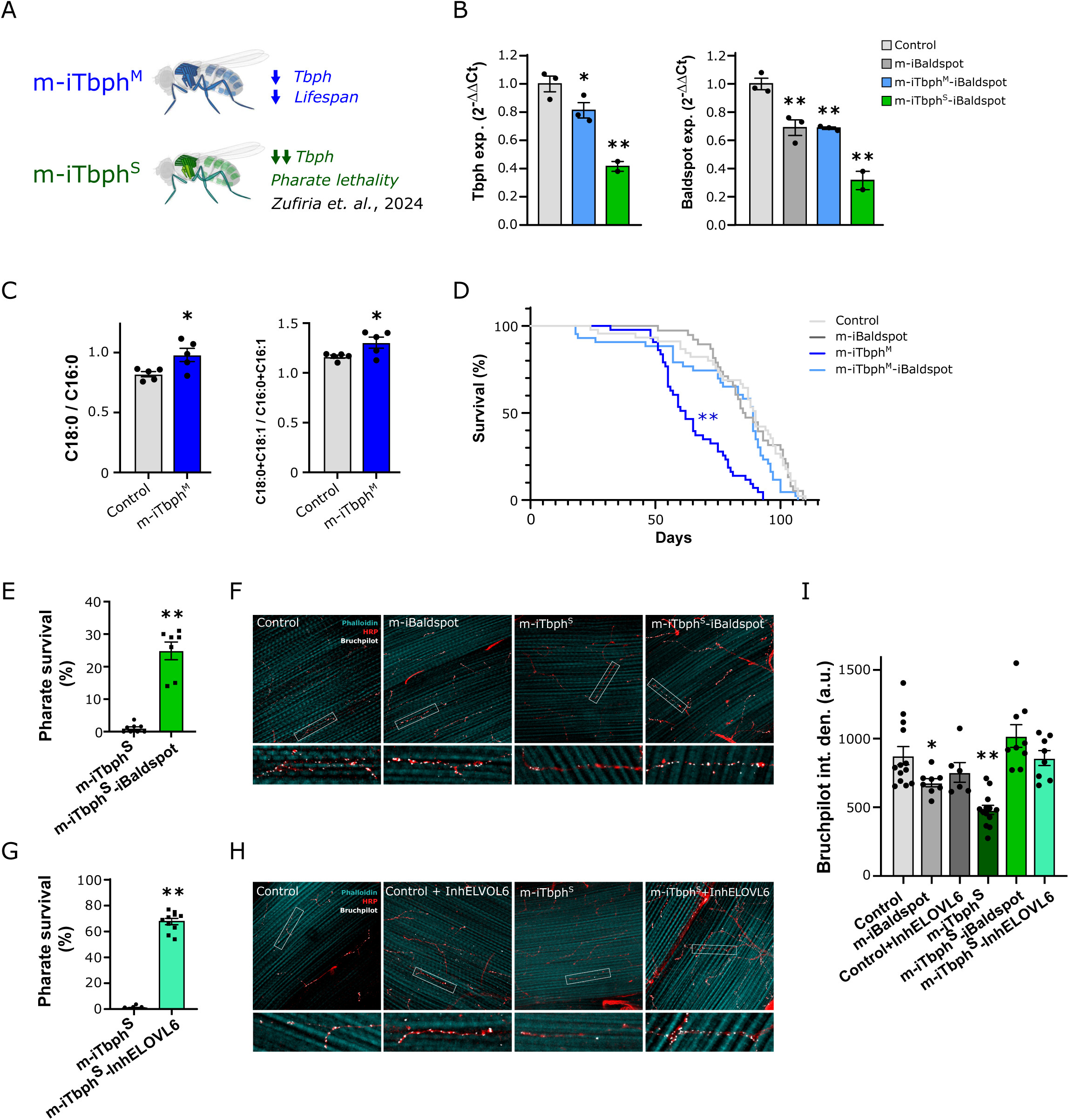
Targeting ELOVL6 activity rescues metabolic reprogramming, survival, and neuromuscular synaptic integrity in *Tbph* loss-of-function models. **(A–B)** Characterization of *Drosophila* models with moderate (iTbphM) and severe (iTbphS) muscle-specific knockdown of *Tbph*, exhibiting reduced lifespan and complete pharate lethality, respectively. **(C)** Metabolic validation. Lipidomic profiling of thoracic muscles reveals that *Tbph* depletion recapitulates the human metabolic signature, characterized by significantly elevated C18:0/C16:0 elongation ratios (p < 0.01). **(D–E)** Genetic rescue of survival. The concomitant knockdown of *Baldspot* (ELOVL6 ortholog) fully restores lifespan in the moderate model **(D)** and rescues approximately 30% of flies from pharate lethality in the severe model **(E)**. **(F–H)** Restoration of synaptic architecture. Confocal imaging of NMJs stained for the active zone marker Bruchpilot (nc-82, magenta) and neuronal membrane (HRP, green). Loss of *Tbph* leads to denervation and active zone loss **(F, H)**, which is rescued by both genetic ablation of *Baldspot* **(F)** and pharmacological treatment with a selective ELOVL6 inhibitor **(H)**. **(G)** Pharmacological rescue. Treatment with the ELOVL6 inhibitor yields a potent, dose-dependent rescue of eclosion in the severe lethal model. **(I)** Quantification of synaptic integrity. Both genetic and pharmacological inhibition of ELOVL6 restore nc-82 signal intensity to control levels, linking metabolic correction to the preservation of the motor unit.

First, we validated the metabolic signature. Thoracic muscles from iTbphM flies showed a significant increase in C16-to-C18 elongation activity (elevated C18:0/C16:0 ratios), mirroring the profile observed in human patients (Figure 3C; Figure S5A). In these Drosophila models, the C18:0/C16:0 ratio was selected as the primary readout of tissue-specific ELOVL6 activity, as it directly captures the rate-limiting step of elongation within the muscle tissue. In the severe iTbphS model, direct comparison was precluded because pharate adults fail to eclose and feed, retaining a “larval-like” lipid profile (high C14:0/C16:0, low C18:0) distinct from adult controls (Figure S5B). Nevertheless, the data strongly link *Tbph* loss-of-function to upregulated long-chain fatty acid elongation.

Genetic ablation of the *Baldspot* completely restored the lifespan of the moderate iTbphM model (Figure 3D). Crucially, *Baldspot* knockdown alone had no adverse effects. In the severe iTbphS model, *Baldspot* inhibition rescued approximately 30% of flies from pharate lethality, allowing successful eclosion (Figure 3E). At the cellular level, this rescue was accompanied by the restoration of NMJ integrity. While iTbphS flies showed a significant loss of the presynaptic active zone protein Bruchpilot (nc-82), concurrent *Baldspot* knockdown fully restored nc-82 signal intensity to control levels (Figures 3F, I).

To validate a pharmacological approach, we treated the severe iTbphS model with a selective indoledione ELOVL6 inhibitor. Computational molecular dynamics simulations predicted that this inhibitor maintains high binding affinity for *Baldspot (Ibarluzea et al., 2025)*. Pharmacological treatment yielded a potent rescue, with up to 70% of flies successfully eclosing (Figure 3G). Subsequent NMJ analysis confirmed that the inhibitor restored presynaptic integrity comparable to genetic knockdown (Figures 3H, I).

### Therapeutic efficacy extends to glial TDP-43 gain-of-function models

Finally, we evaluated the efficacy of ELOVL6 inhibition in a toxic gain-of-function model: flies conditionally expressing human TDP-43 (hTDP-43) in glial cells (Glia-hTDP43) using the REPO-GAL4 driver (Figure 4A). Specific glial expression was confirmed by qPCR (Figure 4B) and immunofluorescence (Figure 4D), resulting in a highly aggressive phenotype characterized by a significantly reduced lifespan (Figure 4C). The rationale for utilizing this model stems from the emerging role of astrocytes as active, non-cell autonomous drivers of motor neuron death. Recent studies have demonstrated that reactive astrocytes (A1 phenotype) undergo a profound metabolic shift, upregulating the synthesis and secretion of long-chain saturated FAs. This neurotoxic lipid profile is primarily driven by the overactivation of ELOVL1, an elongase situated downstream of ELOVL6 that handles the synthesis of saturated lipids with ≥ 20 carbons (Ferrero et al., 2025). Given that ELOVL6 provides the essential C18:0 substrates required for those ELOVL1-mediated saturated FA production, we hypothesized that reactive glial cells exposed to hTDP-43 proteinopathy may exacerbate this elongation axis, promoting a pro-inflammatory and neurotoxic environment.

**Figure 4.**
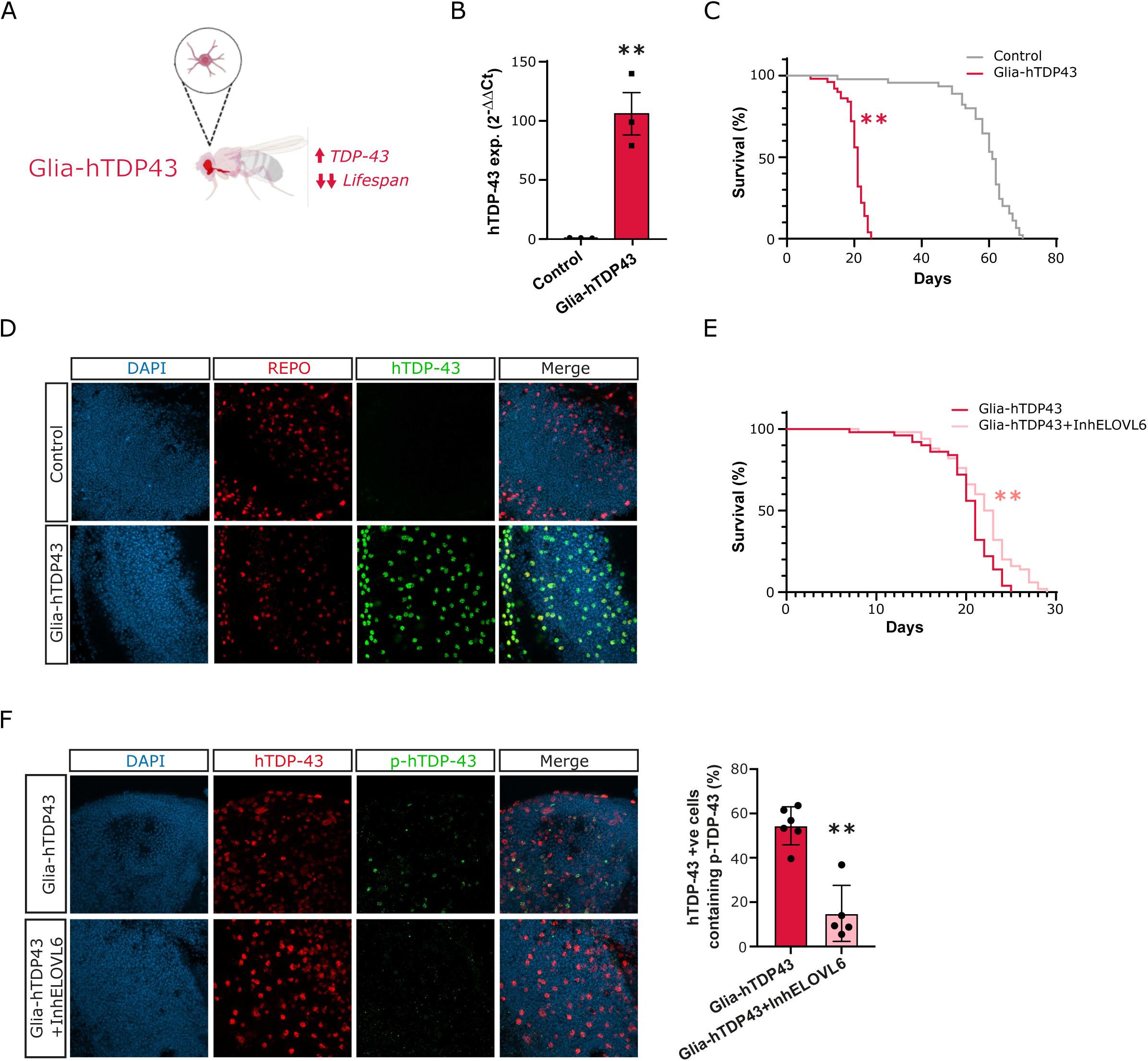
Pharmacological inhibition of ELOVL6 mitigates glial TDP-43 toxicity and reduces pathological protein aggregation. **(A–D)** Validation of the glial gain-of-function model. qPCR **(B)** and immunofluorescence **(D)** confirm the specific expression of human TDP-43 (hTDP-43) within glial cells (Repo-GAL4 driver) of a fly model that has a very shortened lifespan (**C**). **(E)** Therapeutic extension of survival. Kaplan-Meier curves demonstrate that oral administration of the ELOVL6 inhibitor significantly prolongs the lifespan of Glia-hTDP43 flies compared to vehicle-treated controls (p < 0.001), indicating neuroprotection beyond motor neuron-autonomous effects. **(F)** Reduction of proteinopathy. Immunofluorescence analysis of brain tissue reveals the accumulation of pathological phosphorylated TDP-43 (pSer409/410) in the glial model. Notably, treatment with the ELOVL6 inhibitor significantly reduces pTDP-43 levels, suggesting that modulating lipid elongation alleviates the proteostatic stress driving toxic aggregation.

While whole-brain lipidomics did not reveal significant global changes, likely due to the dilution of cell-specific lipid signals within the total brain lipidome (data not shown), the therapeutic impact of ELOVL6 inhibition was robustly demonstrated through functional and pathological readouts. It is well established that reactive astrocytes drive non-cell autonomous neurotoxicity through the targeted synthesis and secretion of long-chain saturated FAs (Ioannou et al., 2019), metabolic shifts that are often undetectable in global tissue assays but are lethal to surrounding neurons. Consequently, our primary evidence for therapeutic efficacy relies on the significant extension of lifespan (Figure 4E) and the reduction of pathological hTDP-43 phosphorylation (pSer409/410) following ELOVL6 inhibition (Figure 4F). These results demonstrate that modulating the glial lipid biosynthetic axis at the enzymatic level is sufficient to mitigate proteinopathy and improve survival.

Taken together, these results demonstrate that ELOVL6 inhibition, whether genetic or pharmacological, potently ameliorates diverse pathological phenotypes (survival, NMJ denervation, and protein aggregation) across both loss-of-function and gain-of-function models, positioning FA elongation as a promising therapeutic target for ALS.

## Discussion

In this study, we demonstrate that the enzyme ELOVL6 functions as a central driver of amyotrophic lateral sclerosis (ALS) rather than a mere bystander in its progression. By integrating clinical lipidomic profiles with functional in vivo validation in Drosophila, we establish that ELOVL6 activity serves as a robust, independent predictor of patient survival and the timeline to critical disease milestones.

ELOVL6 is highly expressed in the liver, adipose tissue and brain, and sits at a critical bottleneck in energy metabolism, catalyzing the elongation of C12–C16 saturated FAs into longer chains (Guillou et al., 2010; Matsuzaka, 2020; Nie et al., 2021; Guo et al., 2025). Our findings suggest that in ALS, this pathway may become hyper-activated, being detrimental to the patient in terms of faster progression rates. This fits with recent evidence showing a fundamental breakdown in how the body handles FA synthesis and desaturation in the spinal cords of ALS patients (Giblin et al., 2024). By forcing the metabolic flux toward long-chain saturated FAs, ELOVL6 likely starves the pool of fats needed for desaturation. This creates a saturation shift that makes cell membranes more rigid and vulnerable to damage and cellular stressors (Volmer et al., 2013; Leamy et al., 2014; Ernst et al., 2018; Giblin et al., 2024; Guerbette et al., 2024; Yang et al., 2025). This metabolic dysfunction has clear clinical consequences. The significant correlation between high ELOVL6 activity and elevated baseline NfL levels further suggests that this metabolic signature is not a static byproduct, but a dynamic marker that mirrors the underlying rate of neuroaxonal injury, effectively acting as a proxy for disease severity. This may be part of a “metabolic trap”: ALS patients often enter a hypermetabolic state where the body shifts toward fat oxidation, and ELOVL6 overactivation may exacerbate the resulting mitochondrial stress and energetic failure. Moreover, ELOVL6 overexpression has been shown to hamper CNS repair and its depletion promotes the induction of a reparative phagocyte phenotype, boosting remyelination in mouse models (Garcia Corrales et al., 2023). Because ELOVL6 is known to regulate insulin sensitivity and lipotoxicity (Matsuzaka et al., 2007; Shimamura et al., 2010; Morcillo et al., 2011), its role in ALS likely represents a systemic failure of metabolic balance.

Validation in *Drosophila* models confirms the causal nature of the ELOVL6-survival relationship observed in our clinical cohorts. By modulating *Baldspot* (the fly ELOVL6 ortholog) in models of TDP-43 proteinopathy, we successfully replicated the pathological metabolic signatures identified in patients, specifically through the elevation of the C18:0/C16:0 ratio, which represents the enzymatic core of the systemic signature, and demonstrated that reversing this imbalance is sufficient to ameliorate disease progression. We observed a complete restoration of lifespan in moderate muscle-specific models and a robust rescue of eclosion rates in severe lethal cases following both genetic and pharmacological intervention. At the cellular level, the restoration of the presynaptic active zone marker nc-82 (Bruchpilot) suggests that inhibiting ELOVL6 preserves the structural integrity of the NMJ, which is widely regarded as the primary site where distal axonal failure begins in ALS (Moloney et al., 2014). This synaptic preservation aligns with recent evidence demonstrating that medium-chain FAs, the metabolic substrates for ELOVL6, exert independent protective effects on NMJ architecture in *Drosophila* models of ALS (Dunn et al., 2023). Since ELOVL6 acts as the enzymatic bottleneck for the conversion of medium-chain FAs into longer-chain saturated species, its inhibition likely provides a dual benefit: preventing the accumulation of neurotoxic long-chain FAs while simultaneously maintaining a pool of beneficial medium-chain precursors that stabilize the synaptic motor unit.

These results also help explain why certain dietary interventions, like n-3 PUFAs, have shown promise in previous studies (Giblin et al., 2024). Since PUFAs are natural suppressors of ELOVL6 (Matsuzaka et al., 2002), the benefits of these fats likely come from their ability to dampen this elongation pathway. However, our data suggests that direct enzymatic inhibition is a much more powerful strategy than dietary supplements. While diets are often limited by how well they can cross the blood-brain barrier (BBB), pharmacological inhibition targets the saturation problem at its source (Giblin et al., 2024).

Our experiments with a selective indoledione inhibitor are particularly encouraging. This compound, which is inferred to have a high binding affinity for both the human and the *Drosophila* enzymes (Ibarluzea et al., 2025), outperformed genetic knockdown in our most severe disease models. A decisive advantage of this pharmacological approach is its capacity for CNS penetrance; unlike dietary FAs (Giblin et al., 2024), which face significant delivery limitations at the BBB, this class of small-molecule inhibitors is designed for high oral bioavailability and effective BBB crossing (Shimamura et al., 2010). The fact that systemic administration successfully reduced pathological TDP-43 phosphorylation directly within glial cells in the brain underscores its therapeutic viability. This suggests that attacking ELOVL6 with a selective drug during key windows of disease progression is not only more effective than permanent genetic changes but also provides a superior alternative to nutritional interventions, positioning ELOVL6 as a promising druggable node for ALS.

The efficacy of ELOVL6 inhibition in rescuing the Glia-hTDP43 phenotype, despite the absence of a global brain lipidomic shift, highlights the importance of cell-specific metabolic nodes. As seen in other models of neurodegeneration, the neurotoxic secretome of glia is driven by a precise upregulation of saturated species like VLSFAs through the ELOVL1 pathway (Guttenplan et al., 2021). Our data suggest that ELOVL6 inhibition acts as a ‘metabolic brake’ within the glia, restricting the supply of substrates for these toxic lipids without requiring a massive alteration of the brain’s structural lipid composition. This is consistent with recent evidence (Giblin et al., 2024) showing that localized enzymatic control of lipid saturation is therapeutically superior to systemic interventions.

Mechanistically, our finding that ELOVL6 inhibition significantly reduces pathological TDP-43 phosphorylation in glia establishes a vital link between lipid metabolism and proteostasis. This neuroprotective effect is likely mediated by a reduction in the metabolic flux towards ELOVL1, an enzyme situated downstream of ELOVL6, thereby limiting the production of neurotoxic very long-chain saturated FAs that reactive astrocytes secrete to drive motor neuron death (Guttenplan et al., 2021). While direct markers of ER stress were not measured in our specific Drosophila models, our results align with established models of lipoapoptosis where saturated lipid imbalances trigger a terminal endoplasmic reticulum (ER) stress response. Specifically, excess saturated lipids have been shown to activate the PERK-eIF2α pathway and subsequent PUMA upregulation, driving neuronal death. In this context, ELOVL6 and its ortholog Baldspot have been previously identified as conserved modifiers of the ER stress response (Palu and Chow, 2018). By targeting ELOVL6, the strategic bottleneck of this biosynthetic axis, we hypothesize that we may effectively alleviate the metabolic burden on the ER and mitigate the pro-inflammatory environment (Garcia Corrales et al., 2023). Furthermore, by reversing the shift toward lipid saturation and restoring membrane fluidity, it is likely that the biophysical capacity of neurons and glia to withstand proteinopathic stress is improved, a mechanism consistent with recent observations across ALS, Parkinson’s, and Alzheimer’s models (Fanning et al., 2019; Hamilton et al., 2022; Giblin et al., 2024).

Collectively, these findings position ELOVL6 as a promising dual-purpose target in ALS. Clinically, systemic ELOVL6 activity provides a quantifiable biomarker to refine patient stratification and prognostic modeling. Therapeutically, it offers a druggable metabolic node that anchors systemic lipid dysregulation to the core mechanisms of TDP-43 proteinopathy, providing a clear roadmap for future disease-modifying interventions.

## Data Availability

All data produced in the present study are available upon reasonable request to the authors

## Abbreviations

ALS: Amyotrophic lateral sclerosis
ELOVL6: Elongation of very long-chain fatty acids protein 6
NfL: Neurofilament light-chain
TDP-43: TAR DNA-binding protein 43
NMJ: Neuromuscular junction
FA: Fatty acid
FVC: Forced Vital Capacity
NIV: Non-invasive ventilation
PEG: Percutaneous endoscopic gastrostomy
RE: Resting energy expenditure
PUFA: Polyunsaturated fatty acid
BMI: Body Mass Index
EMG: Electromyography
FAME: Fatty acid methyl ester
GC-FID: Gas chromatography coupled with a flame ionization detector
Simoa: Single molecule array
LOOCV: Leave-one-out cross-validation
IPC: Inverse probability of censoring
AUC: Area under the ROC curve
CI: Confidence Interval

## Ethics approval and consent to participate

Ethical approval for this study was obtained from the Ethics Committee of Euskadi-Osakidetza (reference number PI2016075); the requirement for ethics approval was not waived. All participants provided written informed consent prior to participation, in accordance with the Declaration of Helsinki. The study was conducted in compliance with Regulation (EU) 2016/679 (General Data Protection Regulation, GDPR).

## Consent for publication

Not applicable.

## Availability of data and materials

The datasets supporting the conclusions of this article (lipid features of human and Drosophila samples) are available from the corresponding author upon reasonable request.

## Competing interests

AA-C declares contracts with the MRC, NIHR and Darby Q17 Rimmer Foundation; consulting fees from Amylyx, Apellis, Biogen, Brainstorm, Clene Therapeutics, Cytokinetics, GenieUs, GSK, Lilly, Mitsubishi Tanabe Pharma, Novartis, OrionPharma, Quralis, Sano, and Sanofi). AAK consulting fees from the UK National Endowment for Science, Technology and the Arts (NESTA).

## Funding

This work was supported by the Study Group in Neuromuscular Diseases (GEEN) of the Spanish Society of Neurology and the Spanish Society of Neurology (to G.F.-E.); the Basque Government (grants #2017222027, #2015111122, #2024333026, and #2025333026 to J.I.R.-S., M.B.R.-L., and F.J.G.-B.; grant #2019222020 to G.G.-C., A.M.-A., and A.L.M.; PhD fellowships #DEDUC15/313 to M.Zuf. and #PRE_2018_1_0095 to J.O.); Bioef (grant #BIO17/ND/023-BD to J.I.R.-S., M.B.R.-L., F.J.G.-B., and I.J.H.); Diputación Foral de Gipuzkoa (#2022-CIEN-000030-01 to J.I.R.-S., M.B.R.-L., and F.J.G.-B.); the Carlos III Health Institute ISCIII/FEDER (grant #PI18/01066 and # PI21/00153 to J.I.R.-S., M.B.R.-L., and F.J.G.-B.); the #StopFugadeCerebros initiative from Roche Farma Spain (to F.J.G.-B.); and IKERBASQUE Basque Foundation for Science (#PP/2022/003 to F.J.G.-B. and #RF/2023/010 to G.G.).

AK is funded by The Motor Neurone Disease Association (1122462), NIHR Maudsley Biomedical Research Centre, ALS Association Milton Safenowitz Research Fellowship (RE19765), the Darby Rimmer MND Foundation, LifeArc (RE23378), MRC (MR/Z505705/1), and the Dementia Consortium (1819242). AAK is supported by the UK Dementia Research Institute through UK DRI Ltd, principally funded by the Medical Research Council. AI and AAC are funded by South London and Maudsley NHS Foundation Trust, MRC (MR/Z505705/1), MND Scotland, Motor Neurone Disease Association, National Institute for Health and Care Research, Spastic Paraplegia Foundation, Rosetrees Trust, Darby Rimmer MND Foundation, the Medical Research Council (UKRI) and Alzheimer’s Research UK.

AA-C is an NIHR Senior Investigator (NIHR202421) and a Visiting Professor at the Perron Institute for Neurological and Translational Science, Australia. This is work was partly supported by an EU Joint Programme - Neurodegenerative Disease Research (JPND) project. The project is supported through the UK MND Research Institute, the following funding organizations under the aegis of JPND - www.jpnd.eu [United Kingdom, Medical Research Council (MR/L501529/1 and MR/R024804/1) and Economic and Social Research Council (ES/L008238/1)] and through the Motor Neurone Disease Association, My Name’5 Doddie Foundation, MND Scotland, LifeArc, Alan Davidson Foundation, and Darby Rimmer Foundation. This study represents independent research part funded by the National Institute for Health Research (NIHR) Biomedical Research Centre at South London and Maudsley NHS Foundation Trust and King’s College London. AAK and AI are visiting senior research fellows at the Perron Institute for Neurological and Translational Science, Australia.

## Authors’ contributions

G.F.-E. and A.Z.-J. contributed equally as first authors. G.F.-E. performed the literature review, acquired and interpreted clinical data, and drafted the manuscript. J.I.R.-S. acquired and interpreted experimental data (fatty acid analysis). R.D., R.F.-T., J.J.P.-A., J.B.E., G.G.-C., A.M.-A., M.Z., and M.P. acquired clinical data. J.R. and M.P. contributed clinical data from the validating and training cohorts, respectively. A.A.K., N.J.A., and A.H. acquired and interpreted NfL data. M.Z., M.B.R.-L., M.Zuf., G.G., and F.J.G.-B. acquired and/or interpreted fatty acid data. A.Z.-J., J.LZ.-E., M.G. performed experiments in fly models. J.O. contributed to data interpretation. S.A.-M., A.A.-C., and I.J.H. revised the manuscript for intellectual content. A.L.M., G.G. and F.J.G.-B. designed and conceptualized the study. F.J.G.-B. analyzed and interpreted data. All authors reviewed and approved the final manuscript.

## Acknowledgements

We thank all the patients with ALS and healthy controls for donating samples to develop this research, supported by the Donostia University Hospital. Thanks to the Basque Biobank, and to Karmele Arnaiz, Natividad Coll and Concepción Galdós for the technical support throughout this study.

