## Supplementary figures and images for "Systemic ELOVL6 activity predicts survival and represents a modifiable target of ALS"

### Supplemental Material

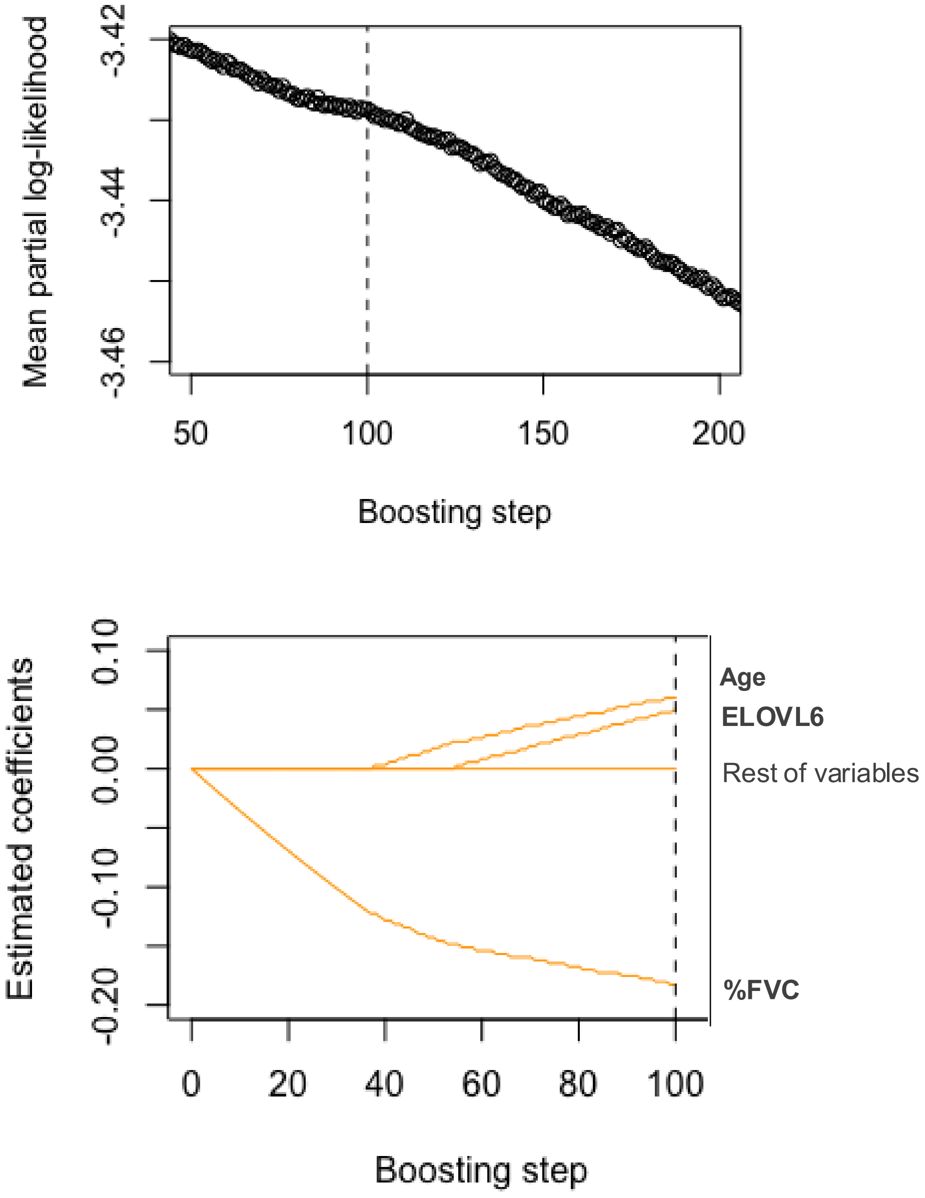

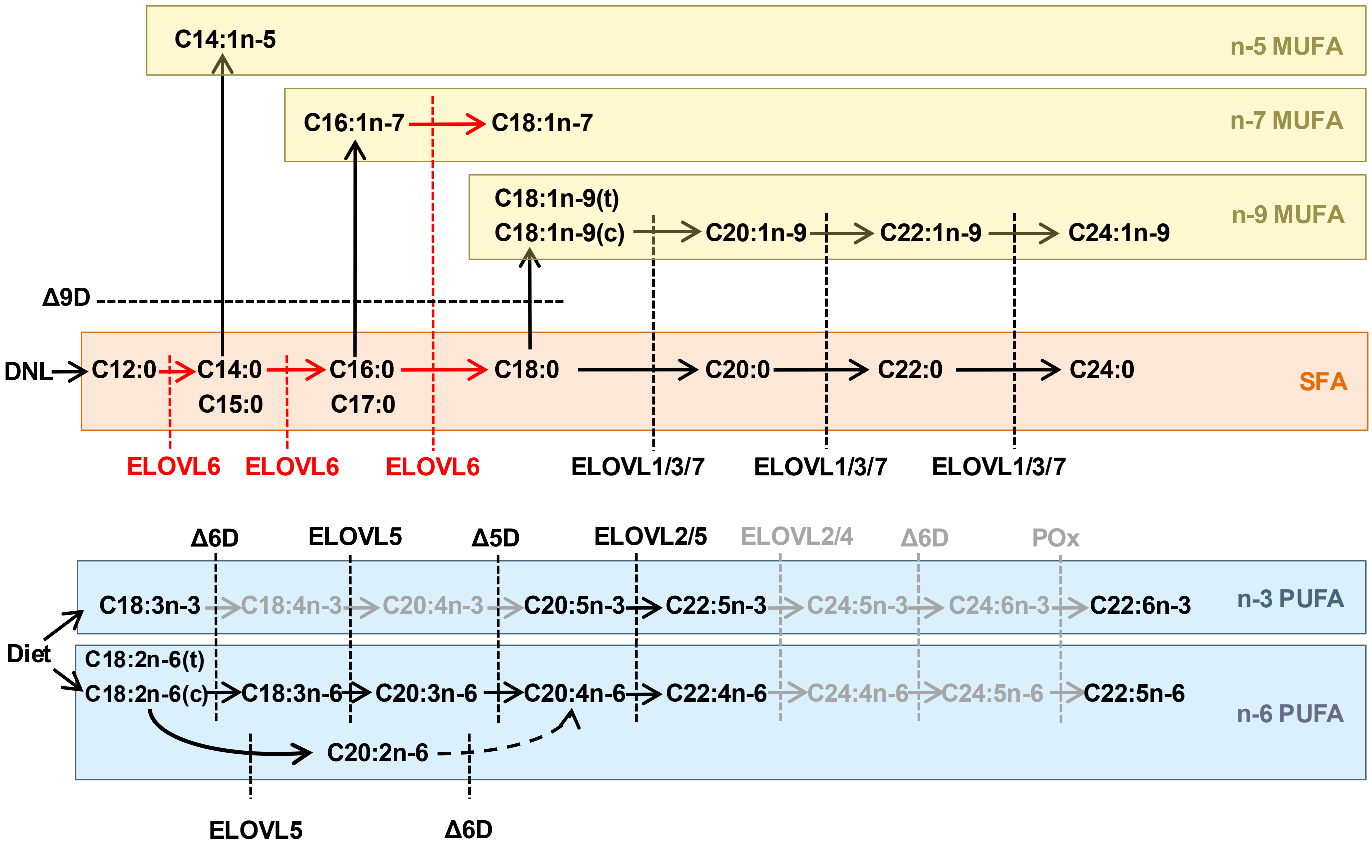

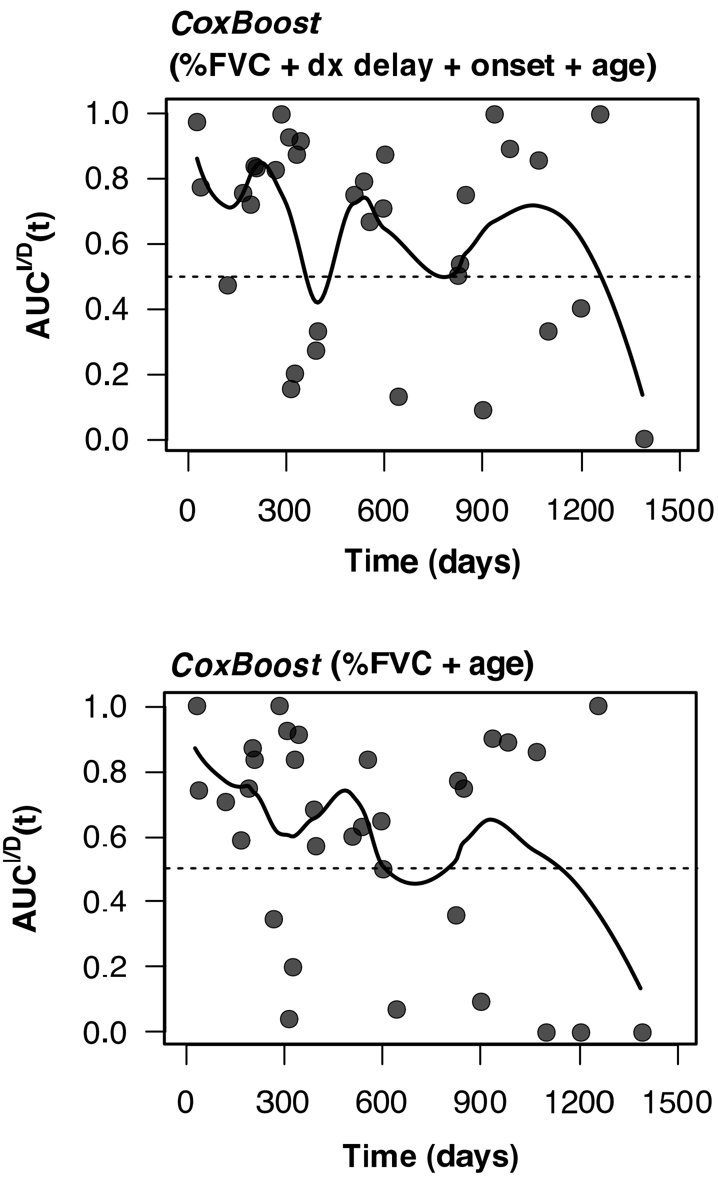

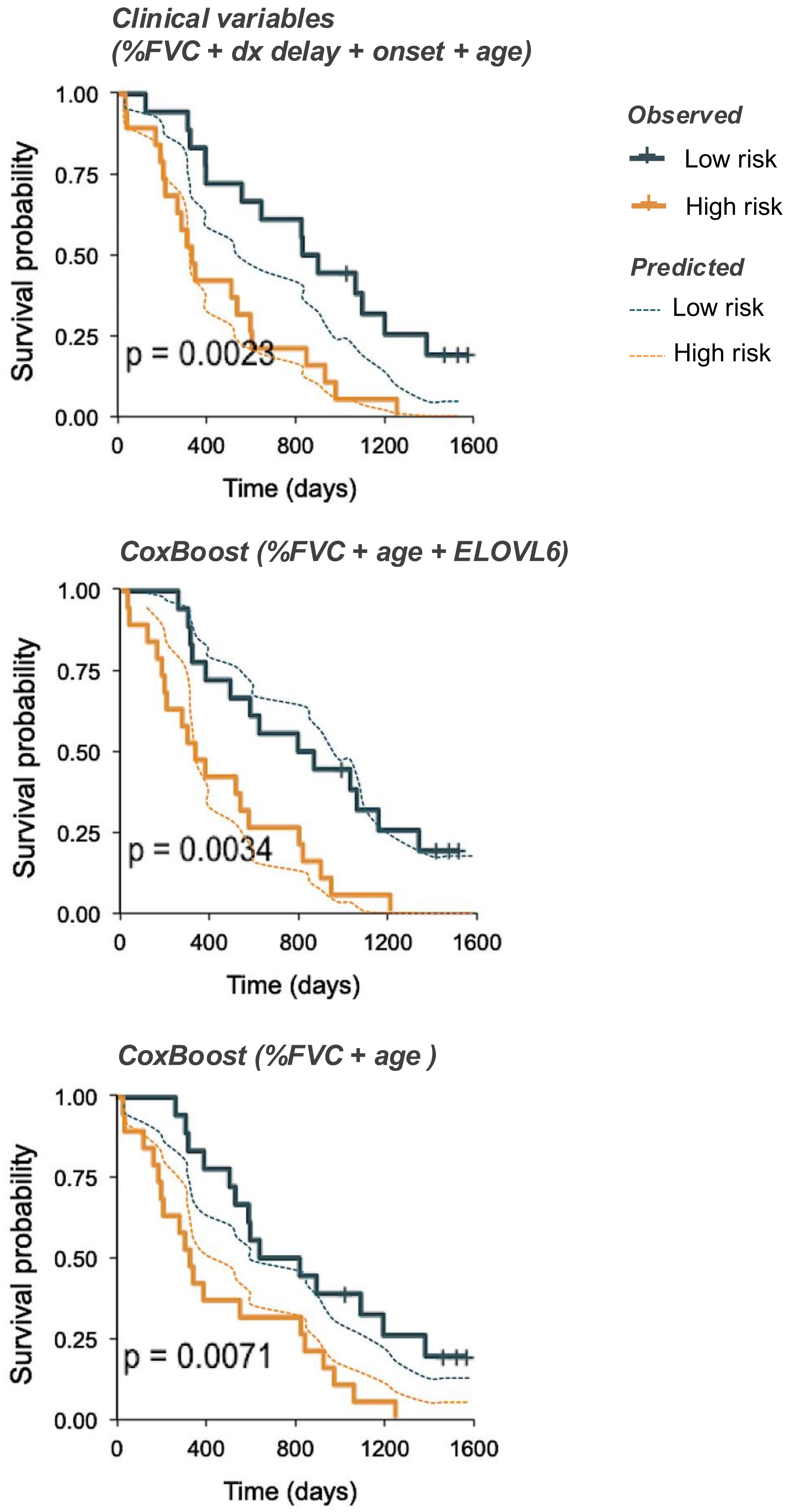

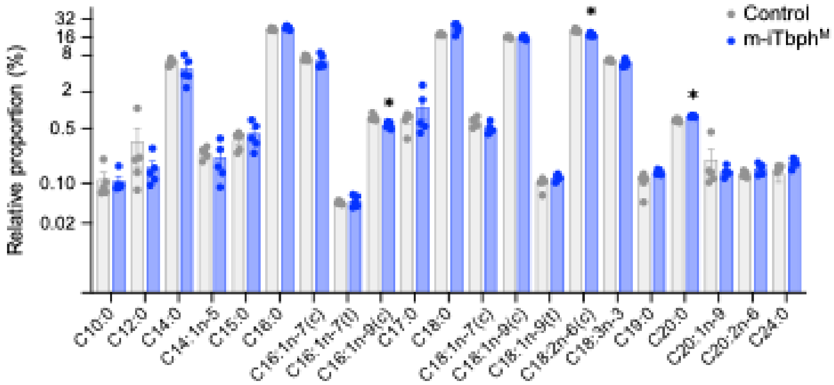

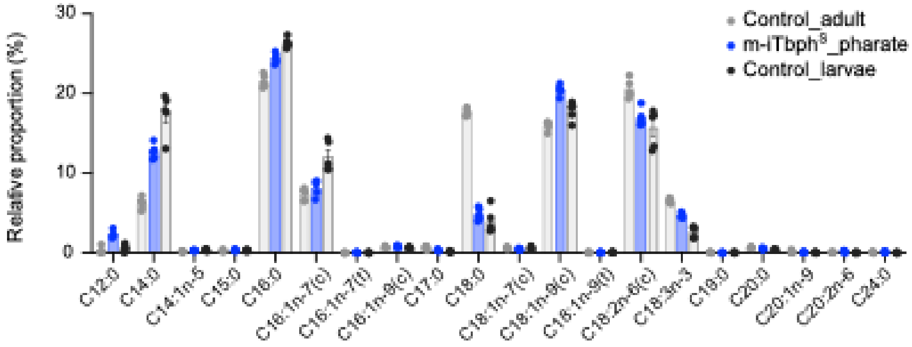
